# The use of complementary and alternative forms of treatment among patients attending Gulu Regional Referral Hospital Mental Health Unit

**DOI:** 10.64898/2026.03.16.26348548

**Authors:** Badriku Kennedy, Avaga Dickens, Outa Paul, Muwanga Ronald, Mpamizo Emmanuel

## Abstract

**Background:** Complementary and Alternative Medicine (CAM) contributes significantly to the utilization of healthcare services in mental health care in sub-Saharan Africa. However, there is limited evidence on the utilization of CAM in the particular setting of post-conflict northern Uganda. This study sought to establish the prevalence, forms, and socio-demographic determinants of CAM use among patients attending the Mental Health Unit at Gulu Regional Referral Hospital (GRRH).

**Methods:** This is a cross-sectional study conducted in a hospital setting from June to August 2025. Convenience sampling was employed to recruit 407 participants. A structured questionnaire was employed for data collection. Data analysis was done using STATA software version 18.0. Descriptive statistics were calculated, and bivariate analysis with Prevalence Ratios (PR) with 95% confidence intervals was employed to determine factors that are significantly associated with the use of CAM.

**Results:** The lifetime prevalence of CAM use was 63.4% (258/407), with 41.3% (168/407) using CAM currently. The most frequent CAM practices used were herbal medicine (50.4%), spiritual practices (33.7%), and traditional medicine (19.8%). For current users, spiritual practices were most frequent (88.7%). The reasons for using CAM were recommendations from others (84.8%) and cultural or religious beliefs (63.4%). Predictors of CAM use were primary education (PR = 1.36, p = 0.017), living in an urban area (PR = 1.23, p = 0.007), separated (PR = 1.39, p = 0.050), and having a mental health disorder for six or more months (PR range = 1.55-1.72). Catholics (PR = 0.72, p = 0.0007) and Protestants (PR = 0.76, p = 0.011) were less likely to use CAM than Born Again Christians.

**Conclusion:** The level of CAM use among patients accessing mental health services in GRRH of northern Uganda is significantly high, while the reporting of CAM use to healthcare providers is remarkably low. This is a challenge that requires urgent attention. Recommendations include integrating the use of CAM into medical practice, developing national policy guidelines on CAM, working in collaboration with traditional/spiritual healers, and conducting public education campaigns.

## INTRODUCTION

Mental health disorders, described as anomalies in cognition, emotion, and mood, have become an increasing concern worldwide [1]. The World Health Organization has recognized mental health disorders as the “pandemic of the 21st century.” There is a large treatment gap, and a large number of people with mental health disorders who are not getting proper health care [1]. Therefore, mental health disorders have implications not only for the individual but also for the family and community [2].

Due to the limitations of conventional medicine, Complementary and Alternative Medicine (CAM) has emerged as a new entity of health practices, properties, and methods that exist independently of conventional medicine [3,4]. CAM refers to natural or holistic healing practices that aim to improve conventional medicine by meeting the unmet health needs and advancing human understanding of medicine [5].

Globally, it has been documented that one out of four individuals suffers from mental health disorders such as depression and anxiety among other mental health disorders [2]. It has been documented that a study conducted on the population of various countries revealed that 3.6% of the world’s population suffering from mental health disorders visit a CAM practitioner, with these figures increasing among individuals residing in high-income countries and those suffering from severe mental health disorders [6]. In the United States, 72% of mental health consumers were found to be using alternative medicine. In Singapore, 6.4% of individuals suffering from mental illness were found to be using CAM, while in Iran, 37.6% of individuals suffering from depression and 68.3% suffering from insomnia were found to be using CAM [8,9].

In fact, over 70% of individuals with mental health concerns initially present to traditional/religious healers in Sub-Saharan Africa, thus causing delays in receiving biological therapies [2,3]. In Nigeria, for example, 87% of patients attending a psychiatric rehabilitation center were found to be using CAM for symptoms, and the main reason for using CAM was cultural/religious beliefs [10]. Traditional medicine is also used by patients with schizophrenia and bipolar disorders in Ghana [11].

The traditional medicinal practices are deeply rooted in the culture of the people of Uganda. These practices are of very important value in the handling of mental health problems. The limited access to conventional mental health services has been recognized to be an important contributing factor to the utilization of traditional, complementary, and alternative medicine (TCAM) services [12,13]. In fact, more than 80% of persons experiencing psychosis have been found to seek conventional biomedical services along with traditional treatment systems. A significant proportion of persons experiencing mental health problems have been found to seek services from traditional faith healers before seeking conventional services [12,14].

In spite of the rising awareness of mental health issues as a public health issue in Uganda, a disconnect still exists with regard to understanding care-seeking behavior, especially with regard to alternative treatment modalities in Northern Uganda and at Gulu Regional Referral Hospital [2]. Such a disconnect may hinder the development of culture-sensitive mental health services, which may affect the pathways to appropriate care for mentally challenged individuals [15, 16].

This study aimed to: (1) determine the prevalence of CAM use among patients attending the Mental Health Unit at Gulu Regional Referral Hospital; (2) identify the types of alternative forms of treatment sought; and (3) determine the socio-demographic factors associated with CAM use.

## METHODS

### Ethics statement

The research was approved by the Gulu University Research Ethics Committee (GUREC) and the Uganda National Council for Science and Technology (UNCST). Permission to conduct the research was sought from the Gulu Regional Referral Hospital Administrator and the Medical Officer In-charge of the Mental Health Department. Informed consent was sought from the participants. The consent forms were read out to the participants because of different levels of literacy among the participants. For participants below 18 years of age or incapable of self-determination, informed consent was sought from the caretakers. The participants were assured of confidentiality and the right to withdraw from the research at any given time. The participants were given refreshments that is soda and biscuits or airtime as a token of appreciation.

### Study design and setting

We used a cross-sectional study design, which was based on the hospital setting and used quantitative research methods. The setting of the research was Gulu Regional Referral Hospital, which is the main referral health facility for Northern Uganda and the neighboring districts, including Amuru, Pader, Lamwo, and Kitgum. It is located at 2.7785 latitude and 32.2980 longitude. The Mental Health Unit at the hospital has outpatient services available every Monday, Tuesday, and Friday, serving approximately 50 patients daily. It also has an inpatient capacity of up to 30 patients. The data was collected from June 17th to August 8th, 2025.

### Study population

The target population comprised outpatients and inpatients undergoing treatment in mental health facilities in Uganda, whereas the accessible population comprised outpatients and inpatients undergoing treatment at the Mental Health Unit of Gulu Regional Referral Hospital, and the sample population comprised patients with a psychiatric diagnosis undergoing treatment at the Mental Health Unit and meeting the inclusion criteria of the study.

### Eligibility criteria

**Inclusion criteria:** Age 2 years and over, since those below 2 years old will be attended to in the pediatric clinics; registered outpatients or inpatients at the Mental Health Unit; and the ability to give informed consent or the presence of a relative/family to give consent.

**Exclusion criteria:** Substantial cognitive impairment or dementia affecting ability to respond accurately; patients too ill to participate; previous involvement in similar CAM studies within the last 6 months; inability to provide informed consent due to profound mental illness or cognitive impairment.

### Sample size determination

Sample size was calculated using the Kish–Leslie formula for prevalence studies: n = (Z² × p(1-p)) / d²

Where:

- n = required sample size

- Z = Z-score corresponding to 95% confidence level (1.96)

- p = estimated prevalence of concurrent use of orthodox biomedical care and traditional treatment systems among individuals diagnosed with psychosis (80%), as reported in previous Ugandan studies [12,14]

- d = margin of error (set at 4%)

Substituting values: n = (3.8416 × 0.16) / 0.0016 = 384

To account for potential non-response, a 6% allowance was added, resulting in a final sample size of 407 participants.

### Sampling method

The method of convenience sampling was used to select mental health patients who were easily accessible at the Mental Health Unit. The researchers interviewed the patients at their regular clinic visits or when they were staying at the inpatient ward. The method was appropriate because it was time-efficient to have an uninterrupted flow of mental health patients receiving active treatment.

### Variables

**Dependent variable:** Complementary and Alternative Medicine (CAM) use (ever used and currently using).

**Independent variables:** Socio-demographic factors (age, sex, education, occupation, residence, marital status, religion, tribe, income); clinical factors (diagnosis, duration of illness).

### Data collection methods and study measures

Data collection involved gathering data from outpatients who attended the Mental Health Unit on Mondays, Tuesdays, and Fridays. The researchers also collected data from inpatients on Wednesdays and Thursdays between the hours of 08:00 and 15:00 when the clinics were in session. The researchers approached the respondents in the waiting area before or after treatment and briefly described the purpose of the research before providing consent forms to those willing to participate in the research. The respondents who agreed to participate in the research were then escorted to a quiet spot to respond to the questionnaires individually.

A structured questionnaire was used to obtain the required demographic, clinical, and CAM-related data. The researchers pre-tested the questionnaires in St. Mary’s Hospital Lacor with 20 respondents (about 5% of the total number of respondents). This pre-test helped to establish the reliability and validity of the questionnaires. About 85% of the pre-test respondents reported that they found the questions in the questionnaires easy to understand. This implied that the questionnaires were clear to respondents. The researchers simplified any question that respondents reported to be too complex. The average time required to complete the questionnaires was about 12 minutes. The reliability of the questionnaires was established through Cronbach’s alpha of 0.86 and test-retest reliability of 0.84 after a week.

### Data quality control measures

- All researchers used the same standardized methods when dealing directly with the patients and when collecting consent and questionnaires.

- The researchers pre-tested the questionnaire among 20 patients in St. Mary’s Hospital Lacor to identify any ambiguities and make corrections.

- The questionnaires were checked on a daily basis to assess their completeness and accuracy.

- The process took place in a quiet environment between 8:00 AM and 3:00 PM.

- The researchers remained motivated through frequent meetings to discuss the process and results.

### Data analysis

For our analysis, we used Microsoft Excel 2021 and STATA 18.0 software. We used descriptive statistics to identify who our participants were and how they used complementary and alternative medicine (CAM). This involved calculating the number and percentage of categorical variables and the mean and standard deviation of quantitative variables such as age.

To identify potential factors associated with the use of complementary and alternative medicine (CAM), we used bivariable analysis to estimate the Prevalence Ratios (PR) of the potential factors using modified Poisson regression. In our case, the Prevalence Ratio of categorical variables was used to compare the prevalence of the use of complementary and alternative medicine (CAM) in the category of interest to the category used as the reference. The 95% Confidence Interval of the Prevalence Ratio is obtained using Cornfield’s Approximate Interval. The level of statistical significance used in our analysis is p < 0.05.

### Community engagement

We worked hand-in-hand with the hospital administration, doctors, nurses, and prominent members of the community, as well as patient advocacy groups, to ensure that the study was conducted in a way that was compatible with what the community needs are. The patients were instrumental in helping shape the direction of the study and the way the results were disseminated to them. We conducted focus groups to create awareness about the study, as well as complementary and alternative medicine and mental health, and then developed a way to collect and address the feedback of the community, so that their needs were addressed appropriately. Once the results were available, we disseminated them to the community, patients, and key stakeholders.

## RESULTS

### Participant characteristics

A total of 407 participants were recruited into the study, and they came from a wide range of sociodemographic backgrounds, as depicted in Table 1 below.

**Table 1:** Sociodemographic characteristics of 407 participants.

| Factor | Description | Frequency(n) | Percentage (%) |
| --- | --- | --- | --- |
| Sex | Male | 191 | 46.9 |
|  | Female | 216 | 53.1 |
| Education | No formal education | 49 | 12.0 |
|  | Primary | 164 | 40.3 |
|  | Secondary | 147 | 36.1 |
|  | University | 35 | 8.6 |
|  | Other | 12 | 3.0 |
| Occupation | Unskilled | 331 | 81.3 |
|  | Semi-professional | 49 | 12.0 |
|  | Professional | 27 | 6.6 |
| Residence | Rural | 188 | 46.2 |
|  | Urban | 219 | 53.8 |
| Marital status | Single | 184 | 45.2 |
|  | Married | 181 | 44.5 |
|  | Other | 1 | 0.3 |
|  | Separated | 4 | 1.0 |
|  | Divorced | 19 | 4.7 |
|  | Widowed | 18 | 4.4 |
| Religion | Born again | 59 | 14.5 |
|  | Catholic | 267 | 65.6 |
|  | Moslem | 7 | 1.7 |
|  | Protestant | 73 | 17.9 |
|  | Other | 1 | 0.3 |
| Income | Low (<300,000 UGX) | 298 | 73.2 |
|  | Middle (300,000 – 500,000 UGX) | 90 | 22.1 |
|  | High (>500,000 UGX) | 19 | 4.7 |

The majority of the participants were women, with 53.1% of them (n=216) compared to men, who comprised 46.9% of the participants (n=191). In terms of education, the majority of the participants were from primary education (40.3%, n=164) or secondary education (36.1%, n=147). The others were from no formal education (12.0%, n=49), university education (8.6%, n=35), or others (3.0%, n=12).

The majority of the participants were unskilled, with 81.3% of them (n=331) compared to 12.0% (n=49) and 6.6% (n=27) of semi-professionals and professionals, respectively. In terms of residence, the majority were from urban areas, with 53.8% of the participants (n=219) compared to 46.2% from rural areas (n=188). The majority were single, with 45.2% of the participants (n=184) compared to 44.5% of the participants who were married (n=181). The others were divorced, widowed, separated, or others, with 4.7%, 4.4%, 1.0%, and 0.3%, respectively.

The majority of the participants were Catholic, with 65.6% of them (n=267) compared to 17.9% and 14.5% of Protestants and Born Again Christians, respectively. The others were Muslim, with 1.7%,

The majority of the participants were from the Acholi tribe, constituting 93.1% of the sample, which equaled 379 participants. The Lango tribe contributed 3.0%, which equaled 12 participants, while the Itesot and Baganda tribes contributed 0.7%, which equaled 3 participants each. The remaining participants were from a variety of tribes, including Alur, Dinka, Jonam, Kakwa, Lugbara, Luo, Madi, Mag, among others (Table 2)

**Table 2:** Tribes of 407 participants.

| **Tribe** | ***Frequency (n)*** | ***Percentage (%)*** |
| --- | --- | --- |
| Acholi | 379 | 93.1 |
| Lango | 12 | 3.0 |
| Itesot | 3 | 0.7 |
| Muganda | 3 | 0.7 |
| Alur | 2 | 0.5 |
| Dinka | 1 | 0.3 |
| Jonam | 1 | 0.3 |
| Kakwa | 1 | 0.3 |
| Lugbara | 1 | 0.3 |
| Luo | 1 | 0.3 |
| Madi | 1 | 0.3 |
| Magwi (Sudanese) | 1 | 0.3 |
| Munyankole | 1 | 0.3 |

### Clinical characteristics

Bipolar affective disorder was the most common diagnosis (37.0%, n=150), followed by epilepsy (30.1%, n=122) and depression (18.2%, n=74). These three diagnoses contributed 85.3% of all reported conditions (Table 3). Other diagnoses included substance use disorder (3.5%, n=14), alcohol-related disorder (3.2%, n=13), schizophrenia (3.0%, n=12), autism (1.5%, n=6), PTSD (0.7%, n=3), somatoform disorder (0.7%, n=3), substance-induced psychosis (0.5%, n=2), and smaller proportions of ADHD, anxiety disorder, cerebral palsy, generalized anxiety disorder, organic psychosis, stress-induced disorder, and suicide attempt (0.3% each).

**Table 3:** Diagnoses of 407 participants.

| **Diagnosis** | **Frequency(n)** | **Percentage (%)** |
| --- | --- | --- |
| Bipolar affective disorder | 150 | 37.0 |
| Epilepsy | 122 | 30.1 |
| Depression | 74 | 18.2 |
| Substance Use Disorder | 14 | 3.5 |
| Alcohol related disorder | 13 | 3.2 |
| Schizophrenia | 12 | 3.0 |
| Autism | 6 | 1.5 |
| PTSD | 3 | 0.7 |
| Somatoform disorder | 3 | 0.7 |
| Substance Induced Psychosis | 2 | 0.5 |
| ADHD | 1 | 0.3 |
| Anxiety Disorder | 1 | 0.3 |
| Cerebral palsy | 1 | 0.3 |
| Generalized Anxiety Disorder | 1 | 0.3 |
| Organic psychosis | 1 | 0.3 |
| Stress induced disorder | 1 | 0.3 |
| Suicide Attempt | 1 | 0.3 |
| **Duration of Mental Illness** |  |  |
| less than 6 months | 37 | 9.1 |
| 6 months to 1 year | 33 | 8.1 |
| 1-2 years | 43 | 10.6 |
| more than 2 years | 294 | 72.2 |

Regarding illness duration, the majority had been diagnosed for more than two years (72.2%, n=294), followed by 1-2 years (10.6%, n=43), less than 6 months (9.1%, n=37), and 6 months to 1 year (8.1%, n=33) as shown in Table 3

The lifetime prevalence of CAM use was 63.4% (258 out of 407 participants) (*fig 1*). Among these 258 lifetime users, all (100%) reported current use of CAM, demonstrating strong ongoing reliance on these practices. The current prevalence of CAM use among all participants was 41.3% (168 out of 407).

**Fig 1:**
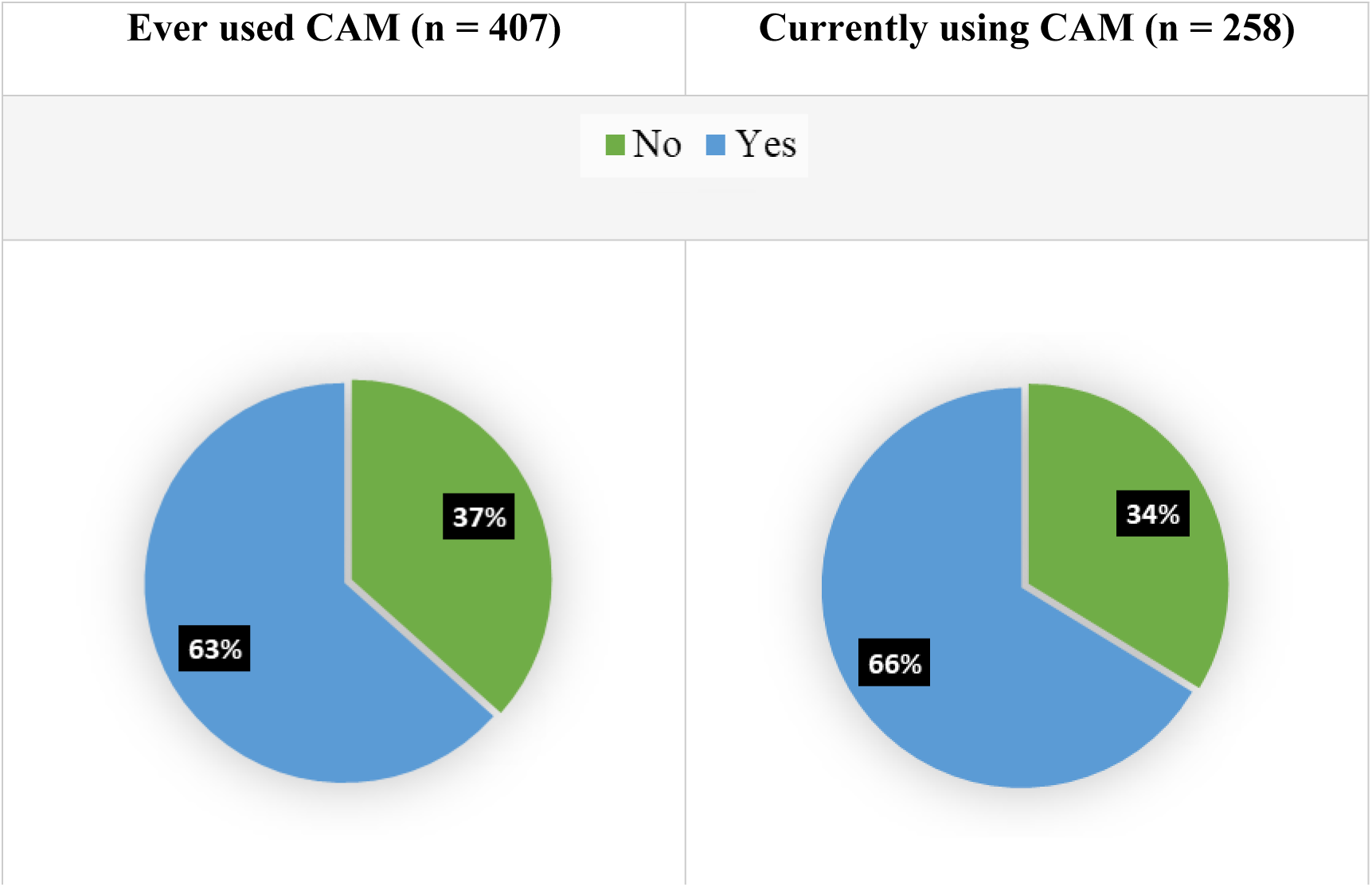
Pattern of CAM use.

Timing and frequency of CAM use: Among lifetime users, 56.1% (n=143) had used CAM for more than two years, 22.8% (n=58) commenced use within the last six months, 11.8% (n=30) started 1-2 years ago, and 9.4% (n=24) began 6-12 months previously. Frequency of use was distributed as follows: occasionally (34.4%, n=87), several times a week (30.4%, n=77), daily (14.2%, n=36), once a week (12.7%, n=32), and rarely (8.3%, n=21) (Table 4).

**Table 4:** Timing of CAM use.

| When they started using CAM (n = 255) | Frequency(n) | Percentage(%) |
| --- | --- | --- |
| More than 2 years | 143 | 56.1 |
| Within the last 6 months | 58 | 22.8 |
| 1-2 years ago | 30 | 11.8 |
| 6–12 months ago | 24 | 9.4 |
| How often they used CAM (n = 253) |  |  |
| Occasionally | 87 | 34.4 |
| Several times a week | 77 | 30.4 |
| Daily | 36 | 14.2 |
| Once a week | 32 | 12.7 |
| Rarely | 21 | 8.3 |

### Types of CAM used

**Lifetime CAM Use:** Among the 258 subjects who have ever used CAM (Table 5), herbal medicine showed the highest frequency of use at 50.4% (n=130), followed by spiritual healing at 33.7% (n=87) and traditional medicine at 19.8% (n=51). Interestingly, considerable cross-over occurred among these CAM modalities because 79.5% of spiritual healing users also used other CAM modalities, 36.4% of traditional medicine users also used herbal medicine, and 24.8% of traditional medicine users also used spiritual healing. Other CAM modalities such as Biodisk, Chinese medicine, etc., were rarely used (0.8%, n=2).

**Table 5:** CAM types ever used among 258 participants.

| **Key** |  |  |  |  |
| --- | --- | --- | --- | --- |
| **Frequency (n)** |  |  |  |  |
| **Percentage (%)** |  |  |  |  |
|  | Herbal | Spiritual | Traditional | Others |
| **Herbal** | 130 |  |  |  |
|  | 50.4% |  |  |  |
| **Spiritual** | 87 | 205 |  |  |
|  | 33.7% | 79.5% |  |  |
| **Traditional** | 51 | 64 | 94 |  |
|  | 19.8% | 24.8% | 36.4% |  |
| **Others** | 0 | 0 | 1 | 2 |
|  | 0.0% | 0.0% | 0.4% | 0.8% |

**Current CAM Use:** Among the 168 current CAM users, spiritual practices were found to be the most dominant at 88.7% (n=149), followed by herbal remedies at 10.1% (n=17), traditional medicine at 0.6% (n=1), and other CAM modalities at 0.6% (n=1). It is interesting to note that the most dominant CAM modality has shifted from herbal medicine to spiritual practices among the current CAM users.

### Reasons for CAM use

From the analysis of the motivations of the 257 participants, it was evident that the most significant motivations for the use of CAM were social recommendations (84.8%, n = 218) and cultural/religious beliefs (63.4%, n = 163) (Table 6). Personal preference was also significant to a lesser extent, as was the availability and costs of CAM. Dissatisfaction with mainstream medicine was also very minimal, at 0.8% (n = 2). Cross-tabulation analysis revealed that spiritual medicine was significantly affected by social recommendations (84.8%) and cultural beliefs (49.4%), and traditional medicine was also significantly affected by cultural beliefs (63.4%).

**Table 6:** Reasons for CAM use among 257 participants.

| Reason | Frequency (n) | Percentage (%) |
| --- | --- | --- |
| Recommendation from others | 218 | 84.8 |
| Cultural/religious beliefs | 163 | 63.4 |
| Personal preference | 33 | 12.8 |
| Availability | 10 | 3.9 |
| Cost | 3 | 1.2 |
| Dissatisfaction with mainstream treatment | 2 | 0.8 |

**Note:** Participants could select multiple reasons, so percentages exceed 100%.

### Disclosure

Importantly, only 23.6% of CAM users had discussed their CAM use with their conventional healthcare provider, highlighting a substantial communication gap. (*Fig 2*)

**Fig 2:**
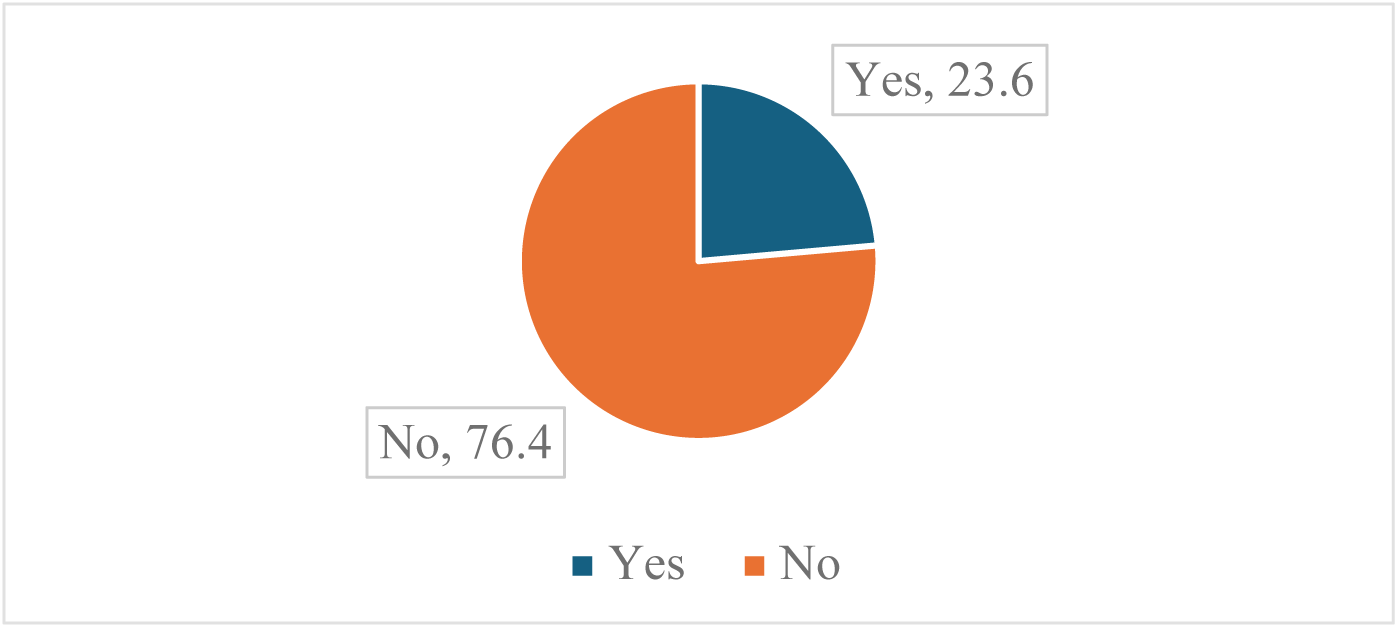
Disclosed to Health Worker.

### Factors associated with CAM use

**Bivariable analysis identified several factors significantly associated with CAM use (Table 4).**

**Education**: Individuals with primary education were more likely to use CAM compared to those with no formal education (PR = 1.36, 95% CI: 1.02–1.83, p = 0.017). Secondary education (PR = 1.19, p = 0.242) and university education (PR = 1.29, p = 0.180) showed non-significant positive associations.

**Occupation:** Semi-professional workers were significantly less likely to use CAM compared to unskilled workers (PR = 0.66, 95% CI: 0.47–0.92, p = 0.003). Professional workers showed a non-significant positive association (PR = 1.25, p = 0.081).

**Residence:** Urban residents were more likely to use CAM than rural residents (PR = 1.23, 95% CI: 1.06–1.44, p = 0.007).

**Marital status:** Separated individuals were more likely to use CAM compared to single individuals (PR = 1.39, 95% CI: 1.10–1.77, p = 0.050). Divorced (PR = 1.67, p = 0.103) and widowed (PR = 1.23, p = 0.237) statuses showed non-significant positive associations.

**Religion:** Catholics (PR = 0.72, 95% CI: 0.62–0.83, p = 0.0007) and Protestants (PR = 0.76, 95% CI: 0.61–0.94, p = 0.011) were significantly less likely to use CAM compared to Born Again Christians. Muslims showed a non-significant negative association (PR = 0.69, p = 0.103).

**Diagnosis duration**: Compared to those diagnosed less than 6 months, CAM use was significantly higher among those diagnosed 6 months to 1 year (PR = 1.72, 95% CI: 1.10–2.70, p = 0.015), 1-2 years (PR = 1.55, 95% CI: 0.98–2.44, p = 0.047), and more than 2 years (PR = 1.62, 95% CI: 1.09–2.41, p = 0.003).

Age (PR = 1.001, p = 0.641), sex (female vs male: PR = 0.96, p = 0.547), and income (middle vs low: PR = 0.95, p = 0.556; high vs low: PR = 1.24, p = 0.171) were not significantly associated with CAM use.

## DISCUSSION

There is limited documented evidence on CAM use among mental health service users at Gulu Regional Referral Hospital (GRRH) in Northern Uganda. This study therefore provides context-specific data from this post-conflict setting. The prevalence of CAM use was high at 63.4% (*Fig 1*), The figure is significantly higher than the global average of 3.6% across 25 countries. [6], It is, however, within the range of 50% to 80% found in other Sub-Saharan Africa countries, where half to four-fifths of those accessing mental health services are using CAM services. [17–19].

It is likely that the high rate of CAM observed in this study is attributed to several factors. First, access to mental health services in post-conflict settings in northern Uganda is limited. This region is characterized by a scarcity of mental health specialists and mental health services in general [20]. Second, the region is characterized by traditional and indigenous beliefs about illness causation and pathogenesis, including spiritual and supernatural causation of illness, which are still prevalent in the region [21,22]. Lastly, the post-conflict setting in northern Uganda is characterized by faith and religion, and faith acts as a coping mechanism in post-conflict settings. Religious leaders are instrumental in psychosocial support in post-conflict settings in northern Uganda [23,24].

Spiritual healing is the most commonly used form of CAM among current users of CAM in post-conflict settings in northern Uganda (87%) (Table 5), although it is also the least used form of CAM when considered in the context of lifetime use of CAM. This indicates that illness becomes chronic and is attributed to spiritual causation and pathogenesis. This is also in agreement with other studies from other Sub-Saharan African settings in which mental illness is attributed to spiritual causation and pathogenesis and in which families seek multiple forms of healing and therapy concurrently [3]. In Nigeria, Ghana, and South Africa, studies have shown that 60-70% of mentally ill persons in these countries used CAM therapies, particularly spiritual healing and herbal medicine. This study contributes to the existing literature on the use of CAM among mental health service users in post-conflict settings in northern Uganda. The results indicate a high rate of 63.4% (*Fig 1*), which is higher than the reported rate of global CAM use of 3.6% in 25 countries worldwide [6]. This is also in agreement with other studies from other Sub-Saharan African settings in which between 50-80% of mental health service users used CAM therapies [17–19].

It is likely that the high rate of CAM observed in this study is attributed to several factors. First, access to mental health services in post-conflict settings in northern Uganda is limited. This region is characterized by a scarcity of mental health specialists and mental health services in general [20]. Second, the region is characterized by traditional and indigenous beliefs about illness causation and pathogenesis, including spiritual and supernatural causation of illness, which are still prevalent in the region [21,22]. Lastly, the post-conflict setting in northern Uganda is characterized by faith and religion, and faith acts as a coping mechanism in post-conflict settings. Religious leaders are instrumental in psychosocial support in post-conflict settings in northern Uganda [23,24].

Spiritual healing is the most commonly used form of CAM among current users of CAM in post-conflict settings in northern Uganda (87%) (Table 5), although it is also the least used form of CAM when considered in the context of lifetime use of CAM. This indicates that illness becomes chronic and is attributed to spiritual causation and pathogenesis. This is also in agreement with other studies from other Sub-Saharan African settings in which mental illness is attributed to spiritual causation and pathogenesis and in which families seek multiple forms of healing and therapy concurrently [3]. In Nigeria, Ghana, and South Africa, studies have shown that 60-70% of mentally ill persons in these countries used CAM therapies, particularly spiritual healing and herbal medicine.

The fact that 84.8% of the subjects were motivated to seek CAM based on social recommendations and 63.4% based on cultural/religious beliefs (Table 6) emphasizes the social aspects of CAM use in this setting. This is supported by other research that found family, friends, and community play a great role in influencing health-seeking behaviors among individuals from collectivistic societies [27]. The low rates of dissatisfaction with conventional treatment (0.8%) suggest that CAM use is complementary rather than healthcare-seeking behavior, which is supported by other research conducted in Uganda indicating that patients may seek healthcare services within both traditional and conventional healthcare systems at the same time [12,14].

Socio-demographic factors were also found to be significantly associated with CAM use. The positive relationship with primary education (PR 1.36) may be attributed to low mental health literacy among individuals with low formal education levels and a strong preference for traditional explanatory models of illness. Similar associations have been found among individuals from Nigeria [10] and Ethiopia [28]. Conversely, semi-professional workers were less likely to seek CAM services than other workers (PR 0.66). This could be attributed to economic stability, better access to conventional healthcare services, and higher mental health literacy among semi-professional workers compared to those from other professions. Similar associations have been found among individuals from Ghana and Kenya, indicating lower rates of CAM use among individuals from formal employment backgrounds [11,29].

The positive relationship with urban dwellers (PR 1.23) (Table 7) contradicts the assumption that CAM is mostly used by individuals from rural areas. It could be attributed to the fact that prayer centers, spiritual ministries, and herbal clinics are common in urban areas, thus making CAM easily accessible to individuals from these areas. Similar associations have been noted among individuals from Kampala, indicating that individuals from urban areas have substantial access to religious and alternative healing centers [30].

**Table 7:**
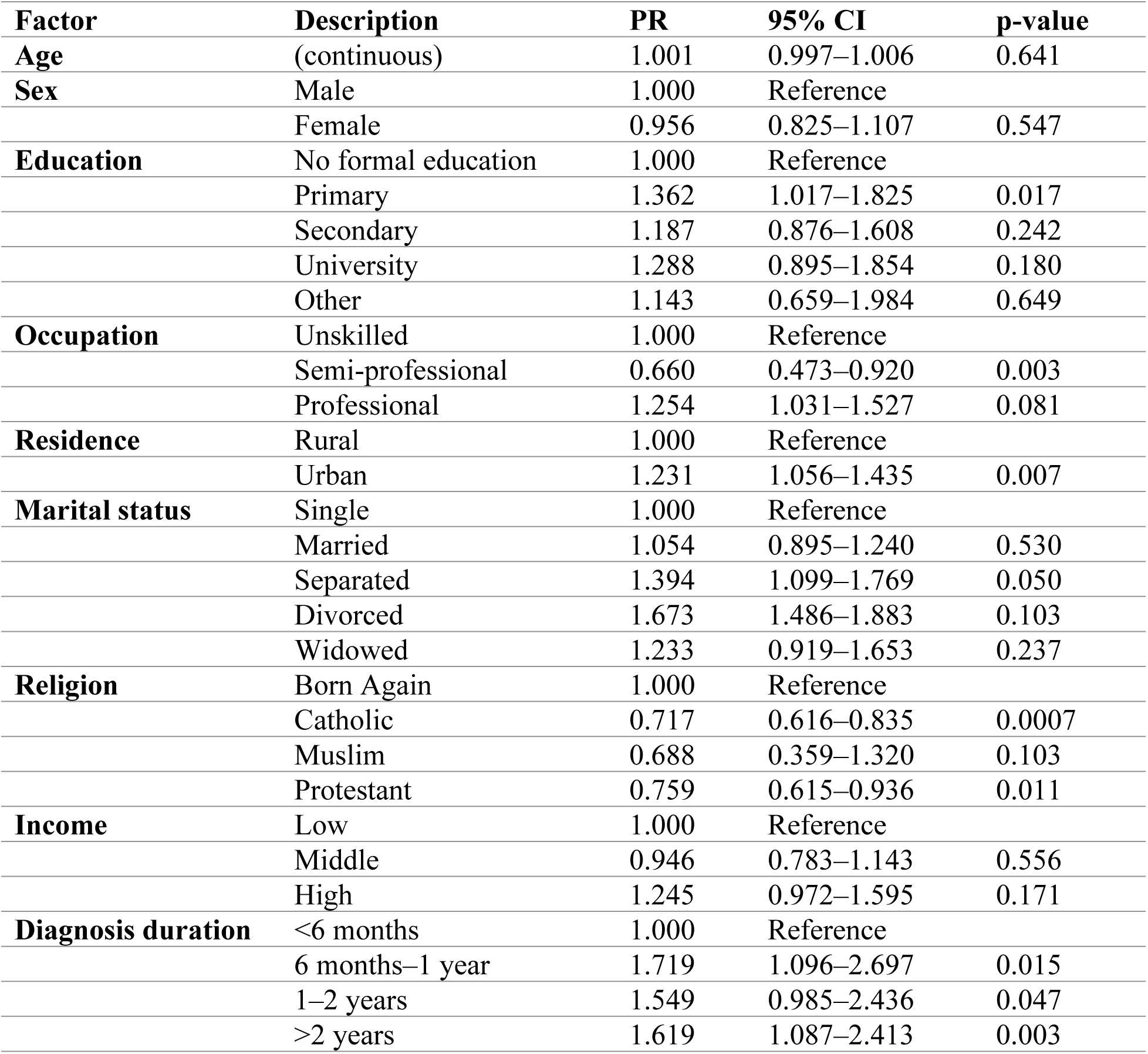
Factors associated with CAM use.

| **Factor** | **Description** | **PR** | **95% CI** | **p-value** |
| --- | --- | --- | --- | --- |
| **Age** | (continuous) | 1.001 | 0.997–1.006 | 0.641 |
| **Sex** | Male | 1.000 | Reference |  |
|  | Female | 0.956 | 0.825–1.107 | 0.547 |
| **Education** | No formal education | 1.000 | Reference |  |
|  | Primary | 1.362 | 1.017–1.825 | 0.017 |
|  | Secondary | 1.187 | 0.876–1.608 | 0.242 |
|  | University | 1.288 | 0.895–1.854 | 0.180 |
|  | Other | 1.143 | 0.659–1.984 | 0.649 |
| **Occupation** | Unskilled | 1.000 | Reference |  |
|  | Semi-professional | 0.660 | 0.473–0.920 | 0.003 |
|  | Professional | 1.254 | 1.031–1.527 | 0.081 |
| **Residence** | Rural | 1.000 | Reference |  |
|  | Urban | 1.231 | 1.056–1.435 | 0.007 |
| **Marital status** | Single | 1.000 | Reference |  |
|  | Married | 1.054 | 0.895–1.240 | 0.530 |
|  | Separated | 1.394 | 1.099–1.769 | 0.050 |
|  | Divorced | 1.673 | 1.486–1.883 | 0.103 |
|  | Widowed | 1.233 | 0.919–1.653 | 0.237 |
| **Religion** | Born Again | 1.000 | Reference |  |
|  | Catholic | 0.717 | 0.616–0.835 | 0.0007 |
|  | Muslim | 0.688 | 0.359–1.320 | 0.103 |
|  | Protestant | 0.759 | 0.615–0.936 | 0.011 |
| **Income** | Low | 1.000 | Reference |  |
|  | Middle | 0.946 | 0.783–1.143 | 0.556 |
|  | High | 1.245 | 0.972–1.595 | 0.171 |
| **Diagnosis duration** | <6 months | 1.000 | Reference |  |
|  | 6 months–1 year | 1.719 | 1.096–2.697 | 0.015 |
|  | 1–2 years | 1.549 | 0.985–2.436 | 0.047 |
|  | >2 years | 1.619 | 1.087–2.413 | 0.003 |

The strong association between longer illness duration and CAM use (PR = 1.55–1.72) is consistent with studies from Pakistan, Iran, India, and Nigeria [8,31–33]. This suggests that with increasing chronicity, patients may experience treatment fatigue, persistent symptomatology, or perceived inadequacy of biomedical interventions, leading them to explore alternative options.

Religious affiliation significantly influenced CAM use, with Born Again Christians showing higher utilization than Catholics and Protestants. Pentecostal and Evangelical practices emphasizing prayer, deliverance, and spiritual healing align closely with CAM modalities [34]. This mirrors findings from Nigeria, Ghana, and South Africa, where CAM use was significantly higher among Pentecostal and charismatic Christians [11,26,31].

Notably, age, sex, and income were not significantly associated with CAM use, suggesting that in post-conflict, resource-scarce settings like northern Uganda, shared cultural beliefs and limited mental health services may override individual demographic factors. This contrasts with high-income contexts where CAM use correlates with female gender, older age, and higher income [35–37].

The low disclosure rate (23.6%) represents a critical finding with significant clinical implications. Non-disclosure of CAM use to healthcare providers creates risks of herb-drug interactions, delayed effective treatment, and fragmented care [7,10]. This communication gap must be addressed through routine clinical inquiry about CAM use in non-judgmental ways.

### Limitations

However, there are a number of things to note. Firstly, it is important to note that since this is a cross-sectional study, it is not possible to establish causative effects. It is also possible that the results obtained might have been skewed in some way in the recollection of the complementary and alternative medicine (CAM) used and the duration of illness. Since the research was conducted in a hospital setting, it is possible that some respondents who used only CAM and never visited conventional medical facilities might have been overlooked. Also, since the majority of the respondents (93.1%) were Acholi, it is possible that the results obtained might not reflect other ethnic groups in Uganda.

Since it is possible that convenience sampling was used in this research, it is likely that the results obtained might not represent any particular group well. Additionally, it is possible that the respondents might have given responses they thought were more acceptable socially.

### Strengths

Despite these challenges, some valuable insights emerge from this understudied research field in northern Uganda. A large sample size (N=407) increases the overall reliability of the study’s results. The quantitative approach enables an exploration of the overall prevalence of CAM and its correlates. The analysis of both lifetime and current CAM use provides a more detailed understanding of its usage. Pre-testing and testing for reliability of the questionnaire increased the overall quality of data collected.

### Implications for practice and policy

From what we have observed, some practical takeaways are obvious. First, at GRRH and other such clinics, patients should be routinely asked about the use of CAM during consultation. This will enhance communication and identify any possible interactions between drugs and herbs. Second, models should be developed to integrate mental health professionals, traditional practitioners, and religious leaders to take advantage of existing community networks. Third, mental health education should incorporate the proper use of CAM, emphasizing its responsible use along with conventional treatment and within treatment guidelines. Fourth, guidelines should be developed at the country level to integrate CAM into mental health services, based on WHO recommendations on traditional medicine. Fifth, conducting surveys at GRRH will provide information on how the pattern of CAM usage changes over time.

Future research should be conducted to assess how the pattern of CAM usage changes over time, to explore the personal significance of CAM to individuals and the reasons for its use through qualitative research, and to assess how collaborative models of conventional and traditional medicine work.

## Conclusion

The research revealed that the prevalence of the use of CAM among mental health patients at Gulu Regional Referral Hospital was significant, with 63.4% of the population relying on CAM when faced with an illness or disease. Spiritual healing and herbal medicine were identified as the most used types of CAM. Several factors were identified as contributing to the use of CAM, which included primary education level, separated status, residing in an urban area, longer illness duration, and religious beliefs. The most surprising discovery was that the majority of the population did not disclose the true cause of illness, with only 23.6% of the population being able to share such information. This is an alarming issue that requires urgent attention. The research revealed that there is a need to develop contextual mental health services that are informed by the use of CAM, patient safety, patient disclosure of information regarding illness causes, and collaboration with spiritual healers to develop policy guidelines at the national level.

## Data Availability

Yes, the data will be available on whenever required basis

https://limewire.com/d/xm1zt#ZY5zdbzCFp

## Acknowledgments

The authors would like to express their appreciation to the study participants for their time and contributions. We thank the administration of Gulu Regional Referral Hospital and the staff of the Mental Health Unit for their support and cooperation. We acknowledge Gulu University for institutional support and the Gulu University Research Ethics Committee for ethical review. We are grateful to the community mobilizers and research assistants who contributed to data collection.

## Author Contributions

**Conceptualization:** Badriku Kennedy, Avaga Dickens, Outa Paul, Muwanga Ronald, Mpamizo Emmanuel

**Data curation:** Badriku Kennedy, Avaga Dickens, Outa Paul, Muwanga Ronald

**Formal analysis:** Badriku Kennedy, Avaga Dickens, Outa Paul, Muwanga Ronald

**Funding acquisition:** Not applicable (no funding received)

**Investigation:** Badriku Kennedy, Avaga Dickens, Outa Paul, Muwanga Ronald

**Methodology:** Badriku Kennedy, Avaga Dickens, Outa Paul, Muwanga Ronald, Mpamizo Emmanuel

**Project administration:** Badriku Kennedy

**Resources:** Badriku Kennedy, Avaga Dickens, Outa Paul, Muwanga Ronald

**Supervision:** Dr. Mpamizo Emmanuel

**Validation:** Badriku Kennedy, Avaga Dickens, Outa Paul, Muwanga Ronald

**Visualization:** Badriku Kennedy, Avaga Dickens, Outa Paul, Muwanga Ronald

**Writing – original draft:** Badriku Kennedy, Avaga Dickens, Outa Paul, Muwanga Ronald

**Writing – review & editing:** Badriku Kennedy, Avaga Dickens, Outa Paul, Muwanga Ronald, Dr. Mpamizo Emmanuel

